# Spatio-temporal variation in the uptake of the Human Papilloma Virus (HPV) vaccine among Malawian girls between 2019 and 2024

**DOI:** 10.64898/2026.02.23.26346859

**Authors:** Jessie Jane Khaki, Alinane Linda Nyondo-Mipando, Donnie Mategula, Fatsani Ngwalangwa, James Chirombo, Mike Chisema, Brenda Mhone, Akosua S. Ayisi, James E. Meiring, Emanuele Giorgi, Mavuto Mukaka, Marc Y. R. Henrion, Michael Give Chipeta

## Abstract

**Background:** Malawi has one of the highest incidences and mortality due to cervical cancer, which is caused by the human papillomavirus (HPV). Achieving high HPV vaccination coverage is critical for advancing the World Health Organization (WHO) cervical cancer elimination strategy. This study aims to describe the spatio-temporal uptake of the first and second doses of the HPV vaccine in Malawi and to investigate the covariates associated with the uptake.

**Methods:** We analysed HPV vaccination coverage data from routinely collected administrative data across 28 districts in Malawi from 2019 to 2024. We used spatio-temporal Bayesian models in R-INLA to investigate the association between environmental factors, such as urbanization and climatic conditions, and vaccination uptake.

**Results:** HPV vaccine uptake was 46.83% (95% Credible interval, CrI: 46.52%, 47.21%) for the first dose, and 32.44% (95% CrI: 32.09%, 32.96%) for the second dose, across the study period, with distinctive subnational heterogeneity. A negative relationship was observed between nighttime light intensity and vaccination coverage (first dose: posterior mean: -0.599, (95% CrI: -1.160, -0.040); second dose: posterior mean: -2.164, (95% CrI: -3.415, -0.967)).

**Conclusions:** HPV vaccination uptake in Malawian districts remains below the WHO 90% vaccination target. These findings emphasise the need for decentralised planning to improve coverage. Targeted interventions, mobile outreach programmes, and strengthened community engagement, particularly in urban settings, may help close coverage gaps and accelerate progress toward cervical cancer elimination in Malawi.

## 1. Introduction

Although cervical cancer, which is caused by several cofactors including the human papillomavirus (HPV), is deemed a preventable disease, it is the third most prevalent cancer and the fourth leading cause of cancer-related deaths in 36 countries [1]. In 2020, there were an estimated 604,127 new cases and 341,831 deaths worldwide, corresponding to an age-standardised incidence rate (ASIR) of 13.3 per 100,000 women (95% confidence interval [CI] 13.3–13.3) and a mortality rate of 7.2 per 100,000 women (95% CI 7.2–7.3) [1, 2]. While other countries have experienced a decline in cervical cancer cases, there has been a rapid increase in the incidence of cervical cancer in several countries, with Malawi registering the most rapid increase in new cases [3]. Malawi also has the world’s highest ASIR (67.9 per 100,000 women, 95% CI 65.7–70.1) and mortality rate (28.6 per 100,000 women) of cervical cancer [1, 2].

As part of cervical cancer prevention strategies, the WHO recommends that at least 90% of all adolescent girls be fully vaccinated with the HPV vaccine by the age of 15 [4]. In order to address the burden of cervical cancer, the Government of Malawi launched a national HPV vaccination program in January 2019 [5]. Under this initiative, the HPV vaccine is administered as a two-dose series to girls aged 9 to 14 years [6]. Initially, the HPV vaccine was delivered in schools by the Ministry of Education and was later integrated into the routine Expanded Programme on Immunisation (EPI) [6, 7, 8]. The program primarily targets 9-year-old girls through school-based and community outreach campaigns, complemented by routine services at health facilities [6].

Literature from other countries shows that uptake of the HPV vaccine is attributable to several factors, including both individual-level and environmental variables [9, 10, 11]. Studies further show that uptake of the HPV vaccine varies spatially [12, 13, 14]. For instance, studies in both developed and developing countries have revealed a spatial socioeconomic gradient in access to, and uptake of HPV vaccine with poorer areas having lower access to and uptake of HPV vaccine compared with richer areas [12, 15, 16].

Although several studies on HPV vaccine uptake have been performed in Malawi, they have focussed at a sub-national level, only including individual districts such as Dowa, Salima, Lilongwe and Zomba [6, 17, 18, 19]. Since the introduction of the HPV vaccine in Malawi, there have been no studies examining the geographical variations in the uptake of the HPV vaccination for the entire country.

Spatial studies that identify areas where health care utilization, including uptake of HPV vaccine is low, are important because they allow for focused planning and delivery of interventions [12, 15]. Uptake of the HPV vaccine is expected to exhibit spatial variation because access to immunisation services, health system capacity, and implementation intensity differ across districts [20]. Geographic differences in school enrolment, outreach coverage, and community acceptance of adolescent vaccination may therefore result in spatial clustering of uptake [21]. Understanding the geographical patterns for the uptake of the HPV vaccine in Malawi may, therefore, help to identify districts with low HPV vaccine uptake and guide implementation partners on which areas to focus interventions such as behavioural communication initiatives. This study provides the first nationally-represented district-level evidence on HPV vaccine uptake since its launch in January 2019. The results from the spatial modelling will help to identify coldspots where uptake of the HPV vaccine is low. These results will guide the Malawi Ministry of Health (MOH) in prioritizing which areas for behavioural communication interventions (BCI) to increase the uptake of the HPV vaccine.

The primary objective of this study is to assess the uptake of the first and second doses of the human papillomavirus (HPV) vaccine among adolescent girls in Malawi. Specifically, we aim to (i) estimate the proportion of eligible girls who received at least one and both doses of the HPV vaccine across the 28 districts using routinely collected surveillance data, and (ii) identify district-level factors associated with the uptake of each dose. To achieve this, separate statistical models were fitted for the first and second dose outcomes.

## 2. Methods

### 2.1. Study area

The study was performed in Malawi, a landlocked country in south-east Africa with an area of approximately 118,500 km^2^. Zambia borders Malawi to the west, Tanzania to the north and northeast, and Mozambique to the east, south, and southwest. Malawi lies at a latitude of around 13.25°S and a longitude of 34.30°E. Administratively, Malawi is divided into 28 districts and 3 regions (North, Central, and Southern).

### 2.2. Study population and vaccination data

The WHO states that the most efficient target group for HPV vaccines are young adolescent girls [18]. This study, consequently, was limited to vaccine-eligible females aged 9–14 years, as per the Malawi national cervical cancer strategy [8]. This study included all 28 districts of Malawi. We obtained annual district-level HPV vaccine coverage data for both the first and second doses for girls aged 9 to 14 years from 2019 to 2024 from the Malawi Ministry of Health. The HPV vaccine coverage per district was defined as the proportion of the target population (girls aged 9 to 14 years) who received at least one dose of the vaccine in each year. Full HPV vaccine coverage per district was defined as the proportion of the target population who received both the first and second doses of the vaccine in a year.

Since routinely collected data are prone to errors, such as coverage values exceeding 100% [22], we capped coverage values at 99%. Specifically, 6% of the 168 records for the first dose and 19% of the 140 records for the second dose exceeded 100% and were therefore capped at 99%, consistent with the approaches used in other HPV studies and in routine childhood vaccination coverage analyses [23, 24].

### 2.3. Geospatial covariate selection for HPV vaccination coverage analysis

For our geospatial analysis of the uptake of the HPV vaccine, we assembled a comprehensive set of geospatial covariates from multiple publicly available sources to explain and predict HPV vaccination coverage patterns across Malawi (see Table 1). These covariates served two purposes: explaining spatial variation in coverage and enabling accurate spatial predictions, with the latter being our primary objective.

**Table 1:** Sources and characteristics of spatial covariates used to model HPV vaccine uptake in Malawi.

| Variable | Temporal Resolution | Data Source |
| --- | --- | --- |
| **Socio-economic indicators** |  |  |
| Nighttime lights (NTL) | 2019-2024 | Earth Observation Group [29] |
| **Climate indicators** |  |  |
| Annual mean precipitation | 2019–2024 | TerraClimate [30] |
| Annual maximum temperature | 2019–2024 | TerraClimate [30] |
| **Accessibility proxies** |  |  |
| Distance to nearest road | 2016 | WorldPop [31] |
| **Sexual behaviour proxies** |  |  |
| Proportion of teenage pregnancy | 2015–16, 2024 | 2015-16 DHS data [32] |
| HIV prevalence | 2015–16, 2020-21 | 2015-16 DHS data [32] & 2020-21 MPHIA [33] |
| **Other environmental factors** |  |  |
| Elevation | 2000 | WorldPop [31] |
| Vegetation index | 2019-2024 | MODIS [34, 35] |
| Temporal positioning | Initial period (2019-2020)<br>Intermediate period (2021-2022)<br>Late period (2023-2024) | Computed from data |

Similar covariates as those shown in Table 1 have been widely used in spatial and geostatistical analyses of healthcare accessibility and utilisation in sub-Saharan Africa, including vaccination and maternal and child health outcomes [25, 26, 27, 9]. Nighttime lights and distance to the nearest road were included as proxies for health system access and service delivery capacity [25, 26, 27]. We also considered behavioural and epidemiological indicators (such as teenage pregnancy and HIV prevalence) to capture underlying adolescent sexual and reproductive health contexts that may influence HPV vaccine demand [7, 28]. Environmental covariates were incorporated to account for geographic and ecological factors that may indirectly affect population distribution and service provision [25, 26, 27].

In addition to these covariates, we also extracted several variables from the 2015-16 Demographic and Health Surveys (DHS) programme and the 2015-16 and 2020-2021 Malawi Population-based HIV Impact Assessment (MPHIA) surveys. District-level HIV prevalence and the proportion of teenage pregnancies were included to approximate sexual behaviour patterns within districts. This approach was informed by previous studies showing that HIV status and teenage pregnancy - used as a proxy for early sexual debut - are associated with an increased prevalence of cervical cancer. [36, 37, 38]. In line with previous spatial mapping studies, we assumed that covariates derived from the DHS and MPHIA remained constant over time. This assumption is justified by the nature of survey estimates, which typically reflect conditions over the five years preceding data collection [39]. Table 1 summarises each covariate, its data source, and temporal coverage. All the spatial covariates were extracted at the district level.

For the temporal positioning variable defined in Table 1, we categorised 2019–2020 as the initial period in Malawi based on observed temporal trends. The years 2021–2022 were classified as the intermediate period, while 2023–2024 were considered the late period. We note that for some of these time points, the classifications coincide with Malawi’s COVID-19 pre-pandemic, pandemic and post-pandemic periods [40].

All numeric variables were standardised prior to inclusion in the models. Each variable was transformed to have a mean of zero (0) and a standard deviation of one (1).

Following the approach taken in previous vaccine uptake geospatial studies [25, 26, 27], we first assessed collinearity between the covariates using a non-spatial generalised linear model and by looking at the variance inflation factor and also plotting a correlation matrix [41]. We found that elevation was highly correlated with maximum temperature (see Appendix C). We therefore excluded it from our analysis.

### 2.4. Spatio-temporal model fitting, validation and prediction

Let *i* = 1, 2, …, 28 denote the district under consideration and *t* represent the year, where *t* = 1, 2, …, 6 for the first dose (2019–2024) and *t* = 1, 2, …, 5 for the second dose (2020–2024). Let *N*_*it*_ denote the total number of eligible girls in district *i* in year *t*, and let *p*_*it*_ represent the probability of vaccination in district *i* in year *t*. We modelled the number of girls vaccinated in district *i* at time *t*, denoted *y*_*it*_, out of the total number of eligible girls (*N*_*it*_) as a binomial random variable: *y*_*it*_ ∼ Binomial(*N*_*it*_, *p*_*it*_). The probability of receiving HPV vaccine is, therefore, modelled using a logit model as:

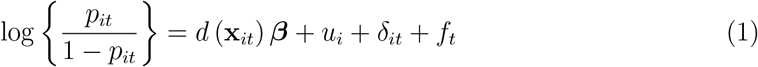

where *β* is a vector of vector of fixed-effect regression coefficients associated with the matrix of spatio-temporal covariates *d* (**x**_*it*_). In equation 1, 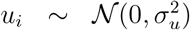 is an unstructured district-level random effect,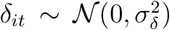 is the unstructured district-year interaction and *f*_*t*_ represents the temporal effect, which was modelled differently across candidate models.

Given evidence from the exploratory analysis of a non-linear pattern in HPV vaccine uptake over time (see Figure A.4 in the Appendix), we explored several non-linear temporal functional forms by specifying latent random walk priors on the time index. A random walk captures gradual, flexible changes over time without imposing a strict functional form, making it suitable for modelling non-linear temporal trends. In our study, it enables the model to adapt to observed fluctuations in HPV vaccine uptake over time.

First, time was modelled as a broken-stick (piecewise linear) function via a first-order random walk (RW1) [42, 43]. This approach assumes the temporal trend consists of distinct linear segments with penalised changes in slope between consecutive time points. In our study, this model can capture abrupt shifts in vaccination rates - for example, rapid increases following the introduction of HPV vaccination into the national immunisation schedule, or sudden declines due to supply disruptions or changes in programme delivery. The RW1 specification is particularly useful for identifying specific time periods when the rate of vaccine uptake accelerated or decelerated.

Building upon the broken-stick model, we next used a penalised spline (P-spline) approximated by a second-order random walk (RW2) [42, 44]. This model penalises second-order differences to induce smoother trends than the RW1 approach, producing a continuously differentiable curve that captures gradual accelerations and decelerations in vaccine uptake. In our study, the P-spline can reveal changes in HPV vaccination coverage driven by sustained factors, such as improvements in health system capacity and increasing community awareness over time.

Finally, we specified a cubic smoothing spline using continuous cubic polynomial basis functions with knots at each time point and a penalised complexity (PC) prior on the smoothing parameter [44, 43]. This model provides the greatest flexibility to capture complex, non-linear temporal trajectories with twice-differentiable smoothness. In terms of HPV vaccine uptake in Malawi, the cubic spline can represent intricate patterns such as sustained exponential growth during early programme expansion phases, plateaus when coverage reaches population limits or multiple inflection points reflecting different policy or implementation phases. The PC prior shrinks the spline toward a linear trend unless supported by evidence, balancing model complexity against parsimony and avoiding overfitting to short-term fluctuations.

Spatial autocorrelation was assessed using the Empirical Bayes Index (EBI), an adaptation of Moran’s I [45]. The empirical Bayes index (EBI) tests indicated negligible residual spatial correlation between districts (see Table B.4). We therefore did not include structured spatial random effects, such as intrinsic conditional autoregressive (ICAR) or Besag–York–Mollié (BYM) priors, in the final models.

Model comparison and selection for both the first and second dose outcomes were performed using the Widely Applicable Information Criterion (WAIC); the model with the lowest WAIC was taken as the preferred specification. Posterior means of of the log-odds ratios were used to predict the uptake of the HPV vaccine over time and space. We also illustrated the uncertainty in the models by visualizing the 95% credible intervals (CrIs) of the predictions. The 95% CrIs were derived from the posterior distribution of the predicted probabilities, taking the 2.5^*th*^ and 97.5^*th*^ percentiles of the simulated posterior samples obtained from the models.

All the analyses were carried out in the INLA package in R [46].

### 2.5. Spatio-temporal prediction

Using the best-fitting model selected for each HPV dose outcome, we carried out spatio-temporal prediction to estimate the number of girls vaccinated in each district and at each time point. Following the approach described by Diggle and Giorgi [47], the first step was to define the predictive target, which in our case was the district-level probability of HPV vaccination, *p*_*it*_, at each time point *t*. For each district *i* and time *t*, we obtained the predicted logit of *p*_*it*_ by combining the estimated fixed effects and random effects from the fitted model.

### 2.6. Model validation (internal)

We assessed the goodness of fit and predictive accuracy of our models through posterior predictive checks by comparing the observed coverage (number of vaccinated girls out of the total targeted) with the model-predicted coverage using Pearson’s correlation [48]. We further evaluated model accuracy using the mean absolute error, root mean square error, and percent bias, with values closer to zero indicating a better model fit [49, 25]. We also illustrated the uncertainty in the models by visualizing the 95% credible intervals of the predictions.

### 2.7. Ethics statement

Permission to use data from the Malawi Ministry of Health (MOH) was requested and received from MOH. Since the data were at aggregated level and hence fully anonymised, it was not necessary to obtain individual consent.

## 3. Results

### 3.1. Description of the sample

Between 2019 and 2024, a combined total of 967,558 girls received the first-dose HPV vaccines and 504,774 girls received the second-dose HPV vaccines across Malawi. HPV vaccine uptake was 46.83% (95% Credible interval, CrI: 46.52%, 47.21%) for the first dose, and 32.44% (95% CrI: 32.09%, 32.96%) for the second dose, across the study period, with distinctive subnational heterogeneity (Figure 1).

**Figure 1.**
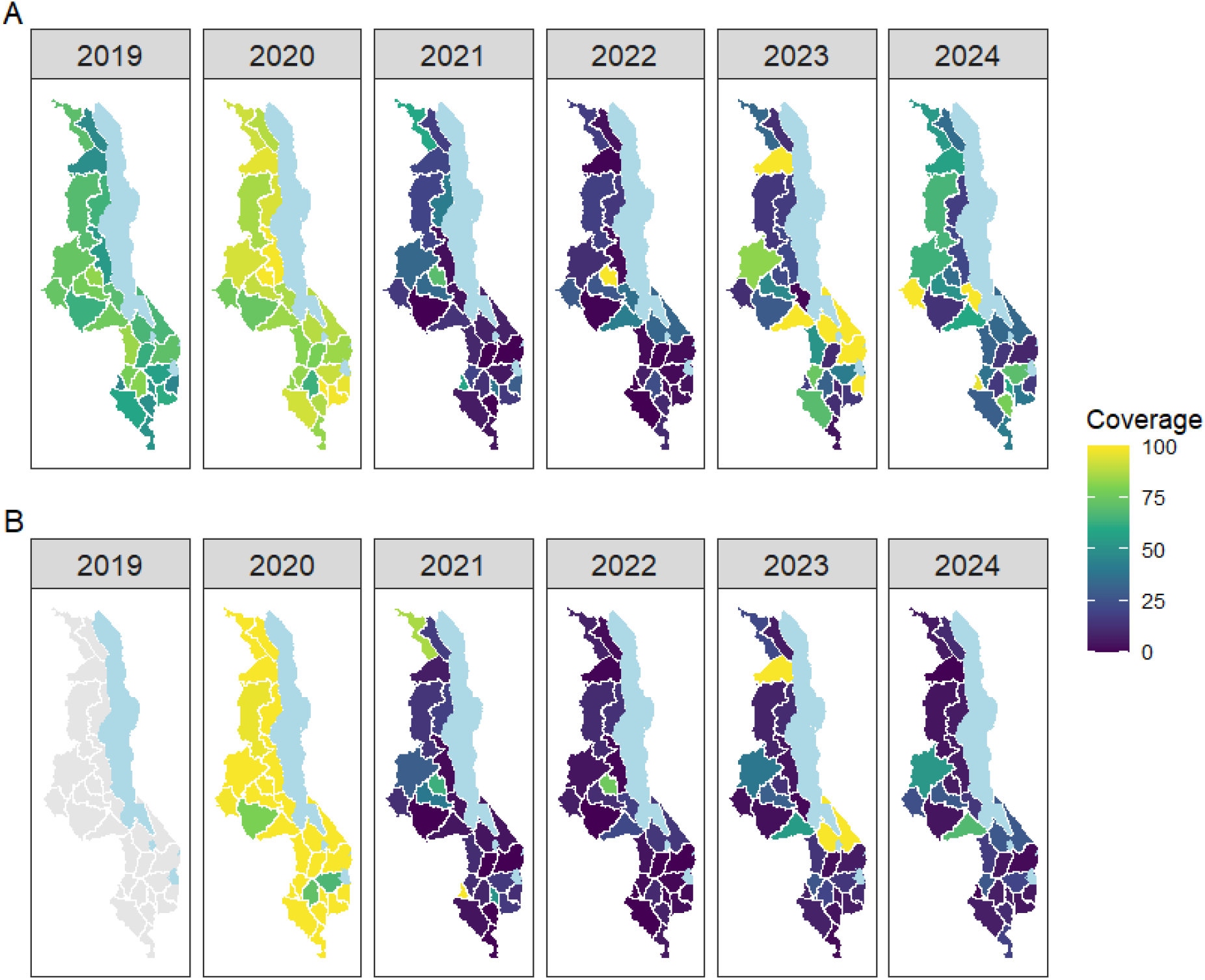
A map showing the tabulated total proportion of young girls who received the first (A) and second (B) doses in each district from 2019 to 2024.

For the first dose (Figure 1, Panel A), coverage in 2019 and 2020 was higher, with greater than half of the districts reaching coverage levels above 75%. However, coverage dropped from 2021 to 2022, when most districts recorded coverage below 40%. The coverage increased again in 2023. Coverage for the second dose (Figure 1, Panel B) was first recorded in 2020, when over two-thirds of districts exceeded 75%. Similar to the first dose of the HPV vaccine, the coverage for the second dose then declined sharply from 2021 onwards, with most districts remaining below 40% through 2024.

### 3.2. Model building and validation

As described in the Methods section, we fitted a range of candidate models. For the first dose outcome, the model with categorical time effects provided the best fit, whereas for the second dose outcome, the model without an explicit temporal function performed best (Table 2). All subsequent results presented in this paper are, therefore, based on these two selected models.

**Table 2:** Selecting a model using WAIC.

| Model | First dose | Second dose |
| --- | --- | --- |
| No time | 1702.62 | **1269.04** |
| Categorical time | **1699.16** | 1559.22 |
| Broken stick | 1702.73 | 1837.53 |
| Penalised spline | 1699.43 | 1829.24 |
| Cubic-like spline | 1700.06 | 1642.60 |

### 3.3. Model validation

In the internal model validation (see Table E.6), the observed and predicted prevalence showed a very high correlation (greater than 95%). Furthermore, the RMSE, MAE, and percent bias for both models were all below 5%. We therefore used the validated models to predict spatio-temporal coverage across the 28 districts and survey years. The full model validation statistics for both doses are reported in Appendix E.

### 3.4. Correlates of the uptake of the HPV vaccine among Malawian girls

Table 3 presents the posterior mean parameter estimates and corresponding 95% CrIs for the final models examining factors associated with the uptake of the HPV vaccine among Malawian girls. For both the first and second dose outcomes, several environmental and temporal factors exhibited notable effects on vaccination coverage, reflected in the magnitude and uncertainty of their posterior distributions.

**Table 3:** Parameter estimates (posterior means of the log odds ratios) and their corresponding 95% credible intervals (CrI) of the final models.

| Parameters | First dose<br>Estimate (95% CrI) | Second dose<br>Estimate (95% CrI) |
| --- | --- | --- |
| *Fixed effects* |  |  |
| Nighttime lights (NTL) | -0.599 (-1.160, -0.040) | -2.164 (-3.415, -0.967) |
| Mean precipitation | -0.319 (-0.832, 0.197) | -2.360 (-3.543, -1.232) |
| Maximum temperature | -0.516 (-1.027, -0.008) | -0.621 (-1.732, 0.483) |
| Distance to nearest road | 0.069 (-0.433, 0.573) | 1.179 (0.077, 2.321) |
| Proportion of teenage pregnancy | -0.407 (-0.967, 0.153) | -1.589 (-2.834, -0.385) |
| HIV prevalence | 0.449 (-0.187, 1.087) | 2.440 (1.098, 3.849) |
| Vegetation index | -0.566 (-1.069, -0.066) | -0.705 (-1.809, 0.378) |
| Temporal positioning |  | NA |
| Intermediate period | *Reference* |  |
| Initial period | 4.297 (3.195, 5.404) |  |
| Late period | 3.827 (2.689, 4.980) |  |
| *Random effects (variance)* |  |  |
| District | $4.54 \times 10^{-5}$<br>($1.17 \times 10^{-5}$, $6.64 \times 10^{-4}$) | $4.53 \times 10^{-5}$<br>($1.15 \times 10^{-5}$, $6.69 \times 10^{-4}$) |
| District-year interaction | 8.35 (6.57, 10.83) | 32.60 (24.71, 44.24) |

NA means not included in the model.

Among the environmental covariates, the posterior distributions indicated that higher nighttime light intensity, a proxy for urbanisation, was associated with a lower probability of vaccine uptake for both dose 1 (posterior mean = –0.599; 95% CrI –1.160, –0.040) and dose 2 (posterior mean = –2.164; 95% CrI -3.415, -0.967). The posterior distribution for maximum temperature also suggested a negative association with uptake of dose 1 (posterior mean = –0.516; 95% CrI –1.027, –0.008). For the second dose, the posterior distribution of mean precipitation also indicated a negative association with vaccine uptake (posterior mean = –2.360; 95% CrI: –3.543, –1.232). Higher vegetation index values showed high posterior support for a negative association with dose 1 (posterior mean = –0.566; 95% CrI –1.069, –0.066), whereas for dose 2 the 95% CrI included zero, indicating weaker posterior evidence of an association.

There was strong posterior evidence that higher district-level HIV prevalence was associated with increased coverage of the second dose of the HPV vaccine (posterior mean = 2.440; 95% CrI: 1.098, 3.849), but little evidence of an association for the first dose. There was also posterior evidence of a negative association between the proportion of teenage pregnancies at the district level and uptake of the second dose of the HPV vaccine, with areas that had higher proportions of teenage pregnancies exhibiting a lower probability of vaccine uptake compared with areas with lower proportions (posterior mean = –1.589; 95% CrI: –2.834, –0.385). The posterior distribution indicated that greater distance to the nearest road was associated with increased uptake of the second dose of the HPV vaccine (posterior mean = 1.179; 95% CrI: 0.077, 2.321).

Compared with the intermediate period, both the initial and late periods showed very strong posterior evidence of higher odds of vaccine uptake. For dose 1, the posterior mean for the initial period was 4.297 (95% CrI 3.195, 5.404), and for the late period 3.827 (95% CrI 2.689, 4.980). The time variable was not included in the second dose outcome model, as shown in Table 2.

Other covariates - including mean precipitation, distance to the nearest road, and the proportion of teenage pregnancies—showed limited posterior support for an association with uptake for the first dose, as their CrIs included zero.

The model random effects also indicated substantial variation of the uptake of the HPV vaccine, especially at the district-yearly level, highlighting the importance of accounting for spatio-temporal heterogeneity in HPV vaccine uptake. There was, however, small evidence of variation at the district level.

### 3.5. Predicted uptake of the HPV vaccine over time

The proportion of young girls receiving the first HPV vaccine dose increased between 2019 and 2020, peaking in 2020 (Figure 2). Coverage then declined in 2021 and 2022. Although uptake improved in 2023 and 2024, it did not return to the levels observed in 2019–2020. For the second HPV vaccine dose, uptake was high in 2020 but declined markedly in 2021 and 2022. Although more girls began receiving the second dose in 2023, coverage declined again in 2024.

**Figure 2.**
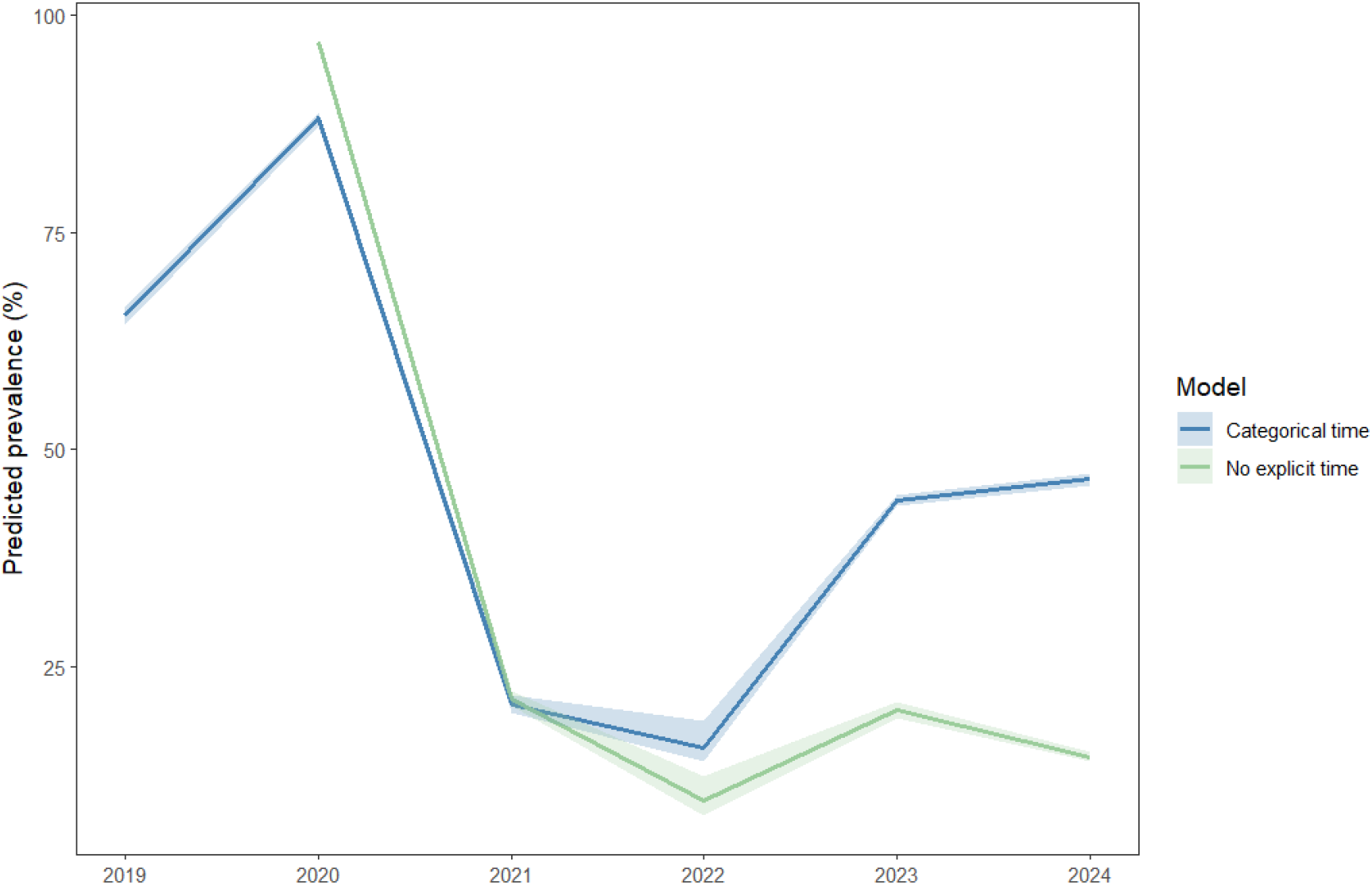
Predicted uptake of the HPV vaccine per year, for the best-fitting models identified in Table 2.

### 3.6. Geographic distribution of HPV vaccine uptake

Due to the lack of residual spatial correlation in the data (see Appendix B.4), we adopted an approach previously used in similar studies to map health outcome data in the absence of spatial dependence [50, 51]. None of the 28 districts met the WHO 90% vaccination target throughout the duration of the study (Figure 3). Areas such as Balaka, Karonga, Nsanje, Nkhotakota, and Mulanje consistently recorded an uptake of less than 45% for both doses. However, a few districts, including Chitipa, Dedza, and Kasungu, achieved the WHO target only once or twice in the 6-year study period, but did not maintain the high coverage. Maps illustrating the uncertainties in the predicted HPV vaccine coverage are shown in Appendix D.

**Figure 3.**
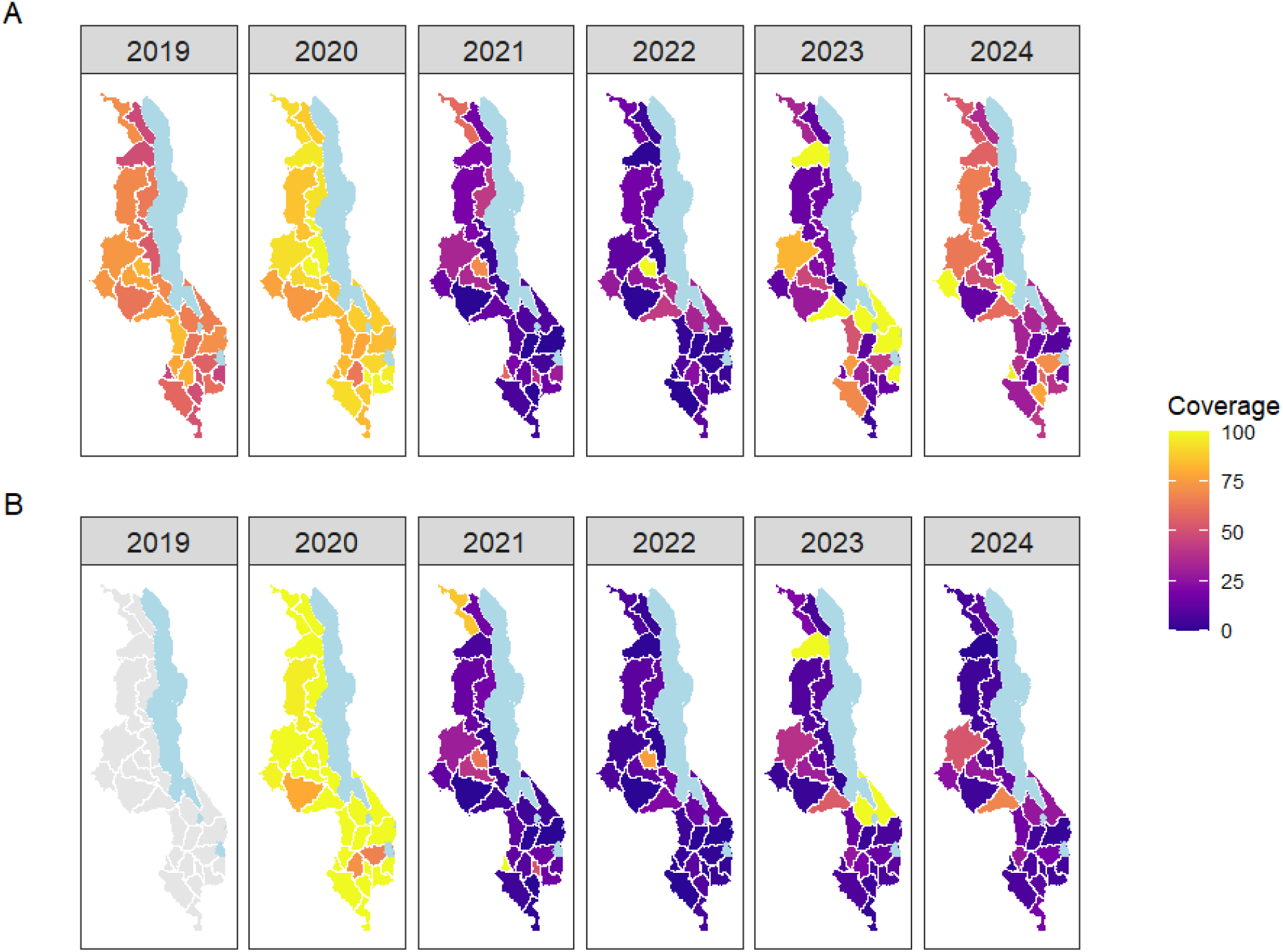
District-level predicted HPV vaccine coverage among adolescent girls in Malawi from 2019 to 2024, for the best-fitting models identified in Table 2 (A = first dose, B = second dose)

## 4. Discussion and Conclusion

### 4.1. Discussion

We present the first national level analysis of the uptake of the HPV vaccine in Malawi since the inception of the HPV vaccine programme in 2019. This study examined environmental, epidemiological, and temporal correlates of the uptake of the HPV vaccine among Malawian adolescent girls, focusing on both the first and second doses. Using spatio-temporal models, we identified associations between HPV vaccine uptake and several contextual factors, supported by their posterior distributions, and predicted geographic and temporal trends. Although there was no residual spatial correlation in the models after adjusting for covariates such as nighttime lights and precipitation, the analysis still revealed substantial geographic heterogeneity in HPV vaccine uptake over time across the 28 districts in Malawi. Patterns of uptake varied across districts from year to year, with no district consistently demonstrating high or low coverage across the study period.

The study found that the average prevalence of HPV vaccination over the study period was 46.83% for the first dose and 32.44% for the second dose. This is lower than the uptake reported in other African countries such as Ethiopia, Tanzania, Uganda and Zimbabwe [52]. These discrepancies might be due to differences in vaccine access and healthcare delivery strategies in these countries [53].

Our findings show a that an increase in nighttime light intensity (NTL), used as a proxy for socioeconomic development, is associated with a lower probability of vaccine uptake. This differs from patterns observed in other countries for both HPV and routine childhood vaccines [9, 27]. Several factors may explain this result. Urban areas in Malawi, particularly townships with high density areas, may experience weaker participation in school-based vaccination programs, or they may have varying attitudes toward HPV vaccination compared to their rural counterparts [54, 55]. Another potential hypothesis is the growing health inequality whereby urban populations face an increasing health burden relative to rural communities [51, 56].

Another finding of the study is that uptake of the second dose increased as the distance to the nearest road increased. suggesting that HPV vaccine coverage is higher in more remote areas. This contrasts with findings from other settings, such as Nigeria and the United States [57, 58]. Nonetheless, it is consistent with our NTL results, which indicate that HPV vaccine uptake in Malawi is higher in rural areas.

The study results also suggest a reduction in HPV vaccine uptake with an increase in the vegetation index, precipitation and temperature with vaccine uptake. These findings are consistent with other studies that show a decline in vaccine uptake with increased precipitation, temperature and vegetation index [59, 60].

Interestingly, an increase in the district-level prevalence of HIV was associated with an increase in HPV vaccine coverage, suggesting that districts with higher HIV burden may have more targeted outreach efforts, potentially due to integrating cervical cancer services into HIV care and other reproductive services [8]. This aligns with previous evidence from different settings, where areas with HIV-related health investments often have better vaccine coverage for other health interventions [28].

Our results also suggest that districts with a high proportion of teenage pregnancies are associated with lower uptake of the HPV vaccine. We hypothesise that in these districts, adolescent girls may have limited access to information on health services, including sexual and reproductive health and HPV vaccination [7]. In addition, girls who are already sexually active or pregnant may be perceived as ineligible for vaccination, or may be excluded in practice, further contributing to lower uptake in high-prevalence districts.

Similar to results reported in other countries, vaccine uptake was higher in the years that did not coincide with the pandemic period [61]. Previous studies in Malawi have hypothesised that school closures and vaccine myths during the COVID-19 pandemic likely contributed to the decline in HPV vaccination coverage during the pandemic [6]. Another study based on case studies from Côte d’Ivoire, Kenya, Liberia, Zambia, and Senegal reported that reduced or suspended HPV vaccination campaigns during the COVID-19 pandemic may have contributed to disruptions in vaccine uptake [62]. We, nonetheless, note that the observed decline in HPV vaccine coverage during the COVID-19 pandemic was not unique to the HPV vaccine but reflected a broader disruption, with reduced uptake observed across the routine immunisation schedule [63, 64].

### 4.2. Conclusion

Our research demonstrates that overall uptake of the first and second doses of the HPV vaccine in Malawi is low compared with WHO targets. The subnational uptake differences highlight the importance of decentralised planning to increase coverage. These findings have important implications for sustaining and expanding HPV vaccination programs in Malawi and comparable LMIC settings as countries work toward meeting the WHO cervical cancer elimination targets.

Our analysis also suggests that vaccine uptake is shaped by a combination of environmental and temporal factors, with urbanization, climatic variability, and pandemic-related disruptions exerting measurable effects. These patterns underscore the need for adaptable and context-specific programmatic responses. Several strategic approaches may help close these gaps. Targeted catch-up campaigns in persistently low-coverage districts could enhance equity in vaccine utilization. Expanding mobile outreach services may also improve vaccine access, particularly during periods of extreme weather or school closures. Integration of the HPV vaccination program with other adolescent health services such as teen clubs offers opportunities to strengthen delivery platforms and improve efficiency [65, 7]. Finally, enhancing community engagement and demand-generation strategies, especially in urban dwellings, may help address vaccine hesitancy and enhance uptake of both doses.

### 4.3. Strengths and limitations of study

A key strength of this study is its use of routinely collected HPV vaccine uptake data, such as HPV vaccine records, to identify geographic areas with low uptake of health services. In the absence of recent national population-level coverage data in Malawi, partly due to funding cuts affecting surveys like DHS, routinely collected data offer practical and scalable alternative for monitoring service delivery and targeting interventions [66, 67]. Additionally, routine data encompasses the entire country, providing comprehensive national coverage that would be difficult to achieve through survey-based approaches alone. To our knowledge, this is the first study to assess HPV vaccine uptake in Malawi using nationally representative data.

The study has several limitations. First, we were unable to incorporate variables such as vaccine hesitancy into our modelling. Second, as is common with routinely collected data, coverage rates occasionally exceeded 100% for reasons discussed in this paper. Despite this data quality challenge, in the absence of survey data such as DHS, our findings remain valuable for program planning and decision-making.

For future research, we recommend triangulating the results from this study with survey-based estimates such as from the Multiple Indicator Cluster Surveys (MICS) should they begin reporting HPV vaccine uptake following the discontinuation of DHS program [68].

This validation would help confirm our findings and provide complementary perspectives on HPV vaccine coverage patterns across Malawi. Future models could explore change-point analysis of the spatio-temporal areal process to identify the precise timing of the COVID-19 pandemic effects on HPV vaccine uptake in Malawi [69].

## Data Availability

All the sources for the data used in this study have been cited in the main manuscript.

https://dhis2.health.gov.mw/

## CRediT authorship contribution statement

Conceptualization: JJK, ALNM, MGC; Data curation: JJK, DM, BM, MC; Formal analysis: JJK; Methodology: JJK, EG, MM, MYRH, MGC; Software: JJK; Validation: JJK; Visualization: JJK; Writing - original draft: JJK; Writing - review and editing: JJK, ALNM, DM, FN, JC, BM, MC, ASA, JEM, EG, MM, MYRH and MGC; Supervision: MYRH and MGC.

## Data availability

All the sources for the data used in this study have been cited in the main manuscript.

## Acknowledgements

The authors would like to thank the Malawi Ministry of Health for providing the HPV vaccine data.

## Appendix A. Raw (tabulated) HPV vaccine coverage trends between 2019 and 2024

Figure A.4 presents the trends in HPV vaccine uptake during the study period. First-dose coverage exceeded 60% in 2019 and 2020, declined to below 20% in 2021 and 2022, and partially recovered in 2023 and 2024. The second dose, introduced in Malawi in 2020, showed a comparable pattern, with coverage falling below 20% in 2021 and 2022, followed by gradual increases in subsequent years.

**Figure A:**
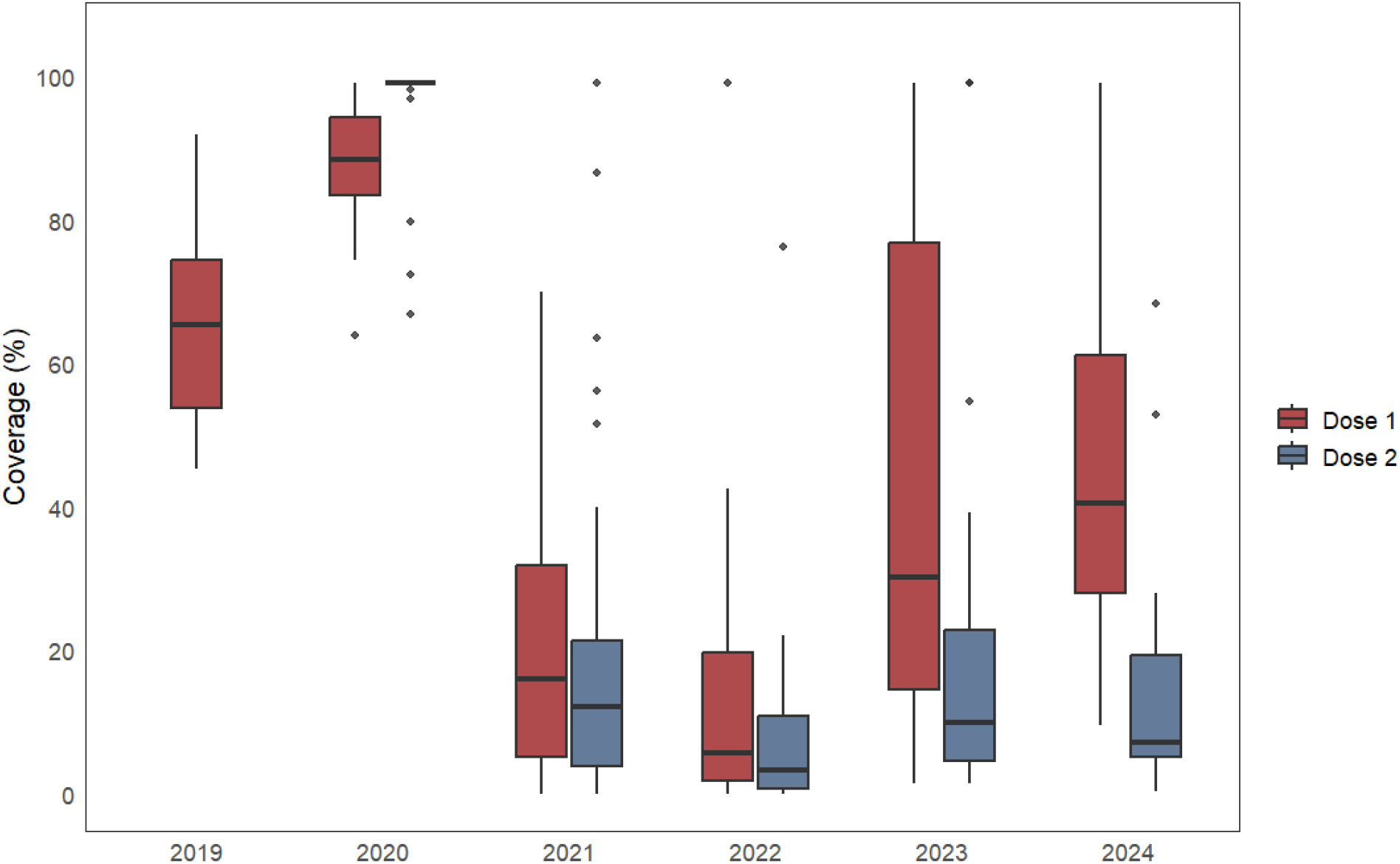
Boxplots showing the temporal trend in the raw uptake of the first dose (2019-2024) and second doses (2020-2024) of the HPV vaccine.

## Appendix B. Assessing the presence of spatial correlation

Spatial autocorrelation was assessed using the Empirical Bayes Index (EBI), an adaptation of Moran’s I. Denoting *m* = 1, 2, …, 28 as the number of districts, *w*_*ij*_ as the (*i, j*)^*th*^ element of the spatial weights matrix *W*, and *Z*_*i*_ as the Empirical Bayes–smoothed estimate of the underlying rate in district *i*, the EBI (*I*_*EB*_) is computed as:

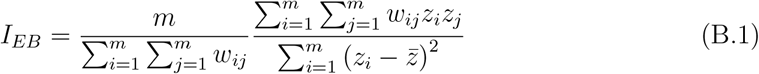

Given that *y*_*i*_ is the number of vaccinated girls and *N*_*i*_ is the number of girls eligible for vaccination in district *i*, the rest of the parameters in B.1 are defined as follows:

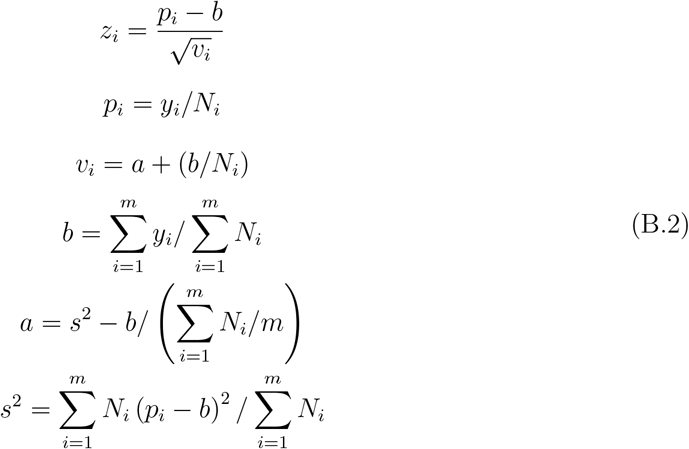

The EBI ranges from -1 to +1, where positive values indicate positive spatial autocorrelation and negative values indicate negative spatial autocorrelation.Using EB-smoothed rates reduces variance instability in sparsely populated areas and limits spurious spatial autocorrelation in Moran’s I. All analyses were conducted using the `EBest`function in the `spdep` package in `R`. Statistical significance was assessed using permutation-based p-values, and p-values below 0.05 were taken as evidence against the null hypothesis of spatial randomness.

Table B.4 summarises the EBI results by year and vaccine dose. For the first HPV vaccine dose, there was evidence of modest but non-random spatial clustering in 2020 (EBI = 0.180, p = 0.032). In 2019, the EBI was small and marginally non-significant (EBI = 0.017, p = 0.071). Between 2021 and 2024, EBI estimates were small and not statistically significant, suggesting no spatial correlation in first-dose coverage.

**Table B.4:**
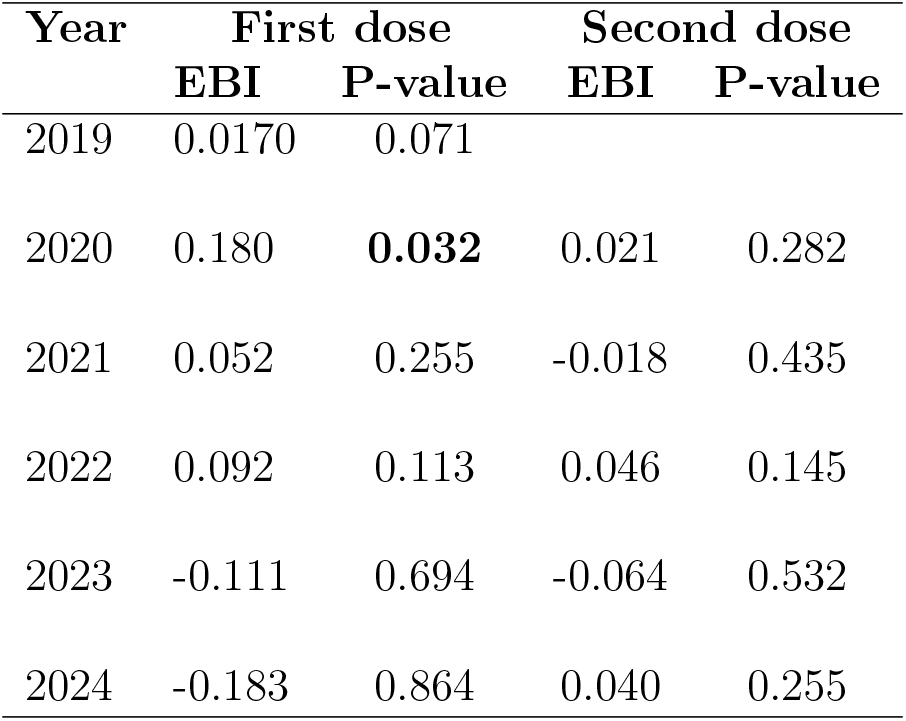
Empirical Bayes Index (EBI) results.

| Year | First dose |  | Second dose |  |
| --- | --- | --- | --- | --- |
|  | EBI | P-value | EBI | P-value |
| 2019 | 0.0170 | 0.071 |  |  |
| 2020 | 0.180 | **0.032** | 0.021 | 0.282 |
| 2021 | 0.052 | 0.255 | -0.018 | 0.435 |
| 2022 | 0.092 | 0.113 | 0.046 | 0.145 |
| 2023 | -0.111 | 0.694 | -0.064 | 0.532 |
| 2024 | -0.183 | 0.864 | 0.040 | 0.255 |

For the second HPV vaccine dose, EBI values were close to zero across all years, and none of the associated p-values were statistically significant. This suggests an absence of detectable global spatial autocorrelation in second-dose coverage during the study period.

## Appendix C. Exploratory analysis and variable processing

We assessed the relationships between the selected covariates using a correlation matrix, as shown in Figure C.5. As illustrated in the figure, elevation was highly correlated (95%) with maximum temperature. Therefore, we excluded elevation from the modelling.

**Figure C.5:**
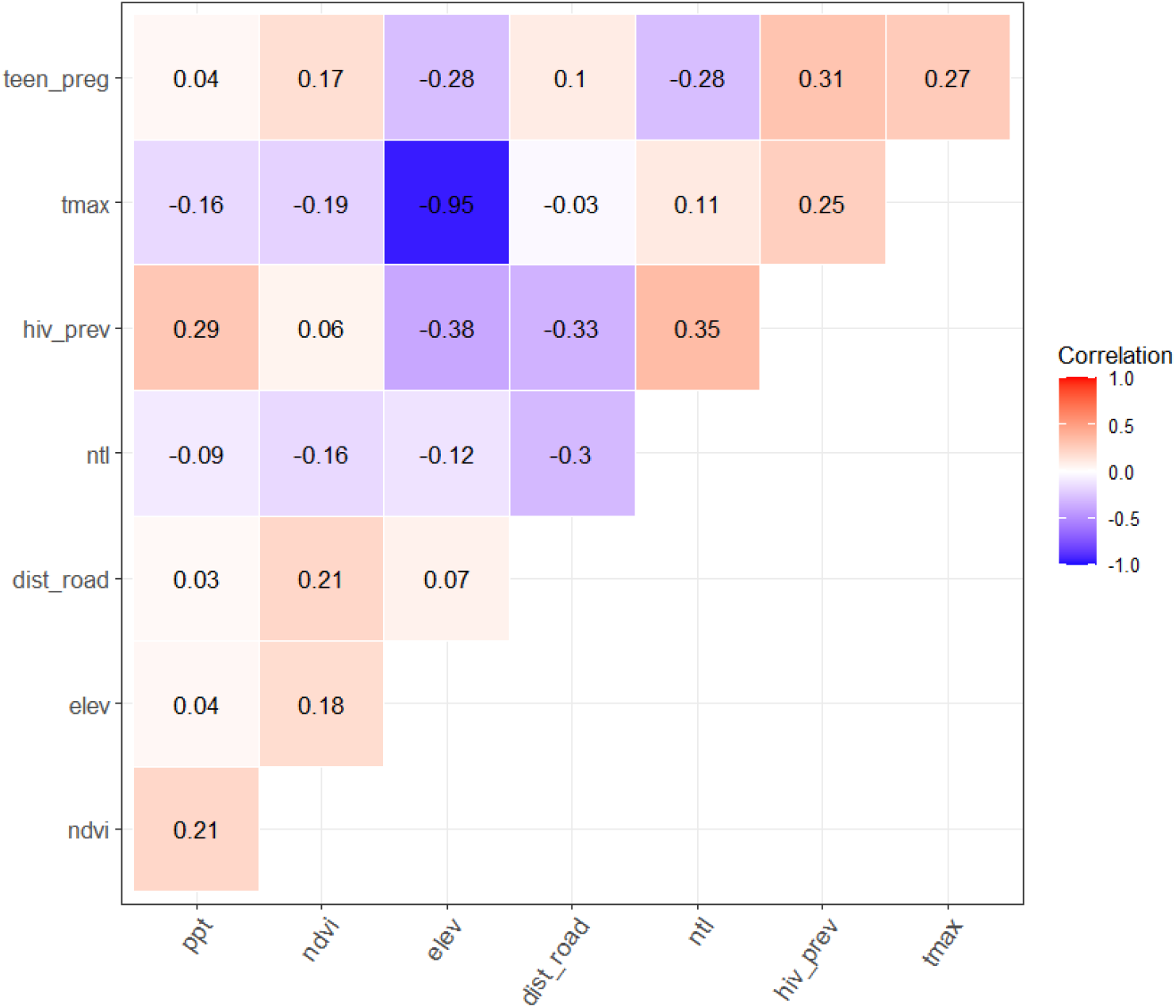
Correlation matrix for covariates.

Table C.5 presents the variance inflation factors (VIF) before and after excluding elevation from the generalised linear model. As shown, the VIF values for maximum temperature and elevation were very high in the initial model. However, after removing elevation, all covariates had VIF values below 3.

**Table C.5:** Variance inflation factors from a generalised linear model.

| Covariate | First dose |  | Second dose |  |
| --- | --- | --- | --- | --- |
|  | A | B | A | B |
| Nighttime lights (NTL) | 2.010 | 1.890 | 1.831 | 1.700 |
| Mean precipitation | 1.811 | 1.772 | 1.786 | 1.756 |
| Maximum temperature | 15.783 | 1.504 | 16.641 | 1.613 |
| Distance to nearest road | 1.364 | 1.350 | 1.422 | 1.410 |
| Proportion of teenage pregnancy | 1.699 | 1.696 | 1.715 | 1.703 |
| HIV prevalence | 2.820 | 2.137 | 2.891 | 2.242 |
| Elevation | 17.722 | Not included | 18.642 | Not included |
| Vegetation index | 1.190 | 1.183 | 1.269 | 1.250 |
| Temporal positioning | 1.573 | 1.320 | 1.743 | 1.400 |

A = Model with elevation, B = Model without elevation.

## Appendix D. Quantifying the uncertainty in model predictions

**Figure D.6:**
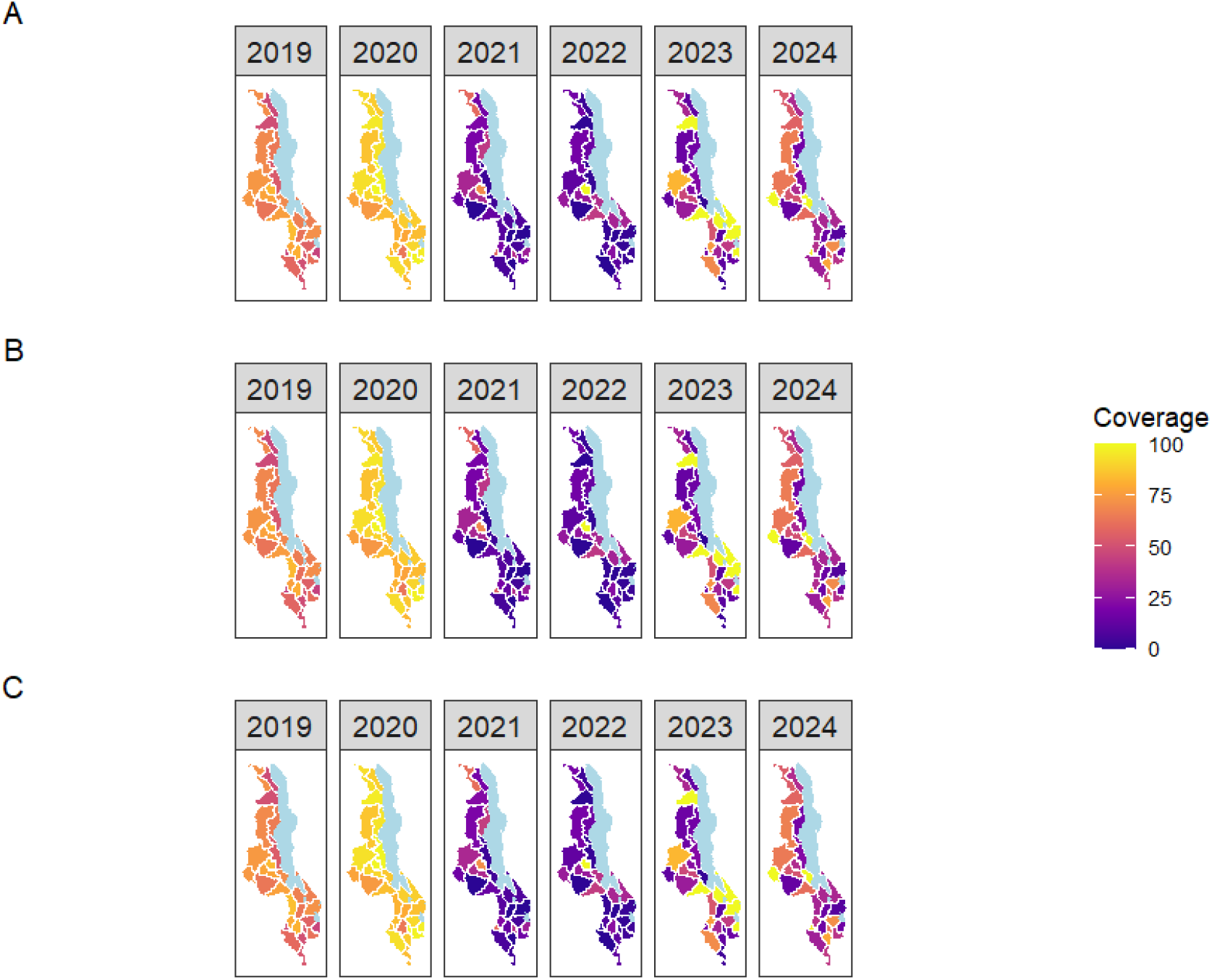
Predicted dose 1 HPV coverage maps: mean predicted prevalence (A) and, lower (B) and upper 95% Credible Intervals (C)

**Figure D.7:**
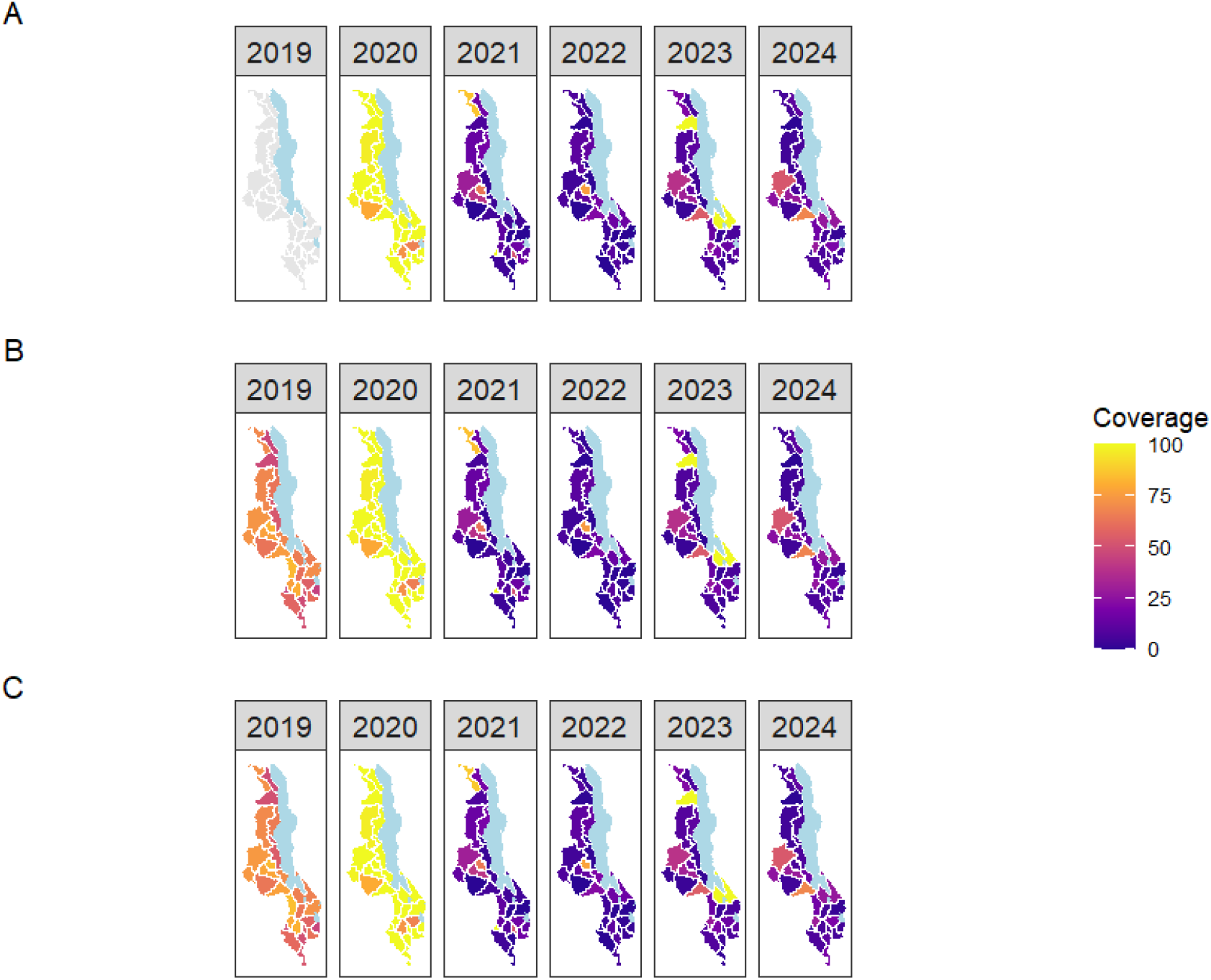
Predicted dose 2 HPV coverage maps: mean predicted prevalence (A) and, lower (B) and upper 95% Credible Intervals (C)

## Appendix E. Model validation results

**Table E.6:** Summary of model validation statistics for the best fitting models.

| Measure | First dose | Second dose |
| --- | --- | --- |
| Correlation coefficient | 0.998 | 0.997 |
| Root mean square error | 0.035 | 0.033 |
| Mean absolute error | 0.006 | 0.004 |
| Percent bias | 0.251 | 0.260 |

## Appendix F. Additional results - Posterior predicted HPV vaccine coverage in non-COVID-19 years

To estimate the posterior predicted coverage of the HPV vaccine in the non-COVID-19 years, we fitted the selected best-fitting models to the dose 1 and dose 2 data after excluding pandemic years. For the first dose, the models were fitted using data from 2019 and 2023–2024; for the second dose, the models were fitted using data from 2023–2024 only. The posterior predicted mean coverage and corresponding 95% credible intervals (CrI) were obtained from the fitted models and are summarised in Table F.7 and Table F.8.

**Table F.7:**
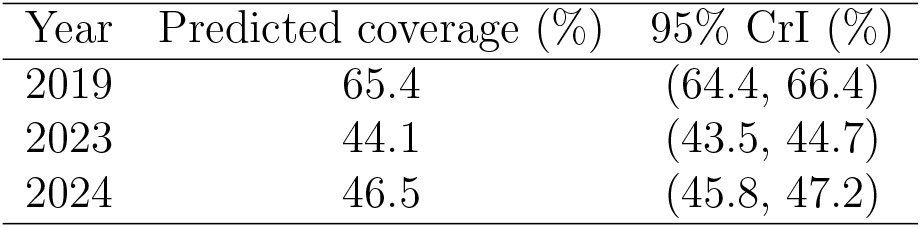
Model-based predicted coverage for Dose 1 by year (non-pandemic years)

| Year | Predicted coverage (%) | 95% CrI (%) |
| --- | --- | --- |
| 2019 | 65.4 | (64.4, 66.4) |
| 2023 | 44.1 | (43.5, 44.7) |
| 2024 | 46.5 | (45.8, 47.2) |

**Table F.8:** Model-based predicted coverage for Dose 2 by year.

| Year | Predicted coverage (%) | 95% CrI |
| --- | --- | --- |
| 2023 | 19.9 | (19.0, 20.9) |
| 2024 | 14.6 | (14.1, 15.1) |

